# Multi-modal recruitment efficiency in ScreenPlus, a large-scale consented pilot NBS program

**DOI:** 10.64898/2026.06.30.26356857

**Authors:** Megan Clarke, Katrina Paleologos, Nicole R. Kelly, Sean M. Bailey, Marjorie Joseph, Gabriel S. Kupchik, Rishi Lumba, Jaya Ganesh, Annemarie Stroustrup, Joseph Orsini, Aaron J. Goldenberg, Melissa P. Wasserstein

## Abstract

ScreenPlus is a consented pilot program that aims to screen 100,000 babies for a panel of rare disorders. Given its size, ScreenPlus provides a unique opportunity to learn about optimal recruitment practices. ScreenPlus’ recruitment strategy includes recruiter-initiated Active and Hybrid modes and parent-initiated Independent mode. Active recruitment occurs in-person at the postpartum bedside, whereas Hybrid recruitment includes other attempt types. In Independent recruitment, parents access online educational and e-consent forms. Analysis of 47,642 completed recruitment profiles from May 2021 through April 2025 showed that Active recruitment was used in 72.2% and had the highest percentage of parents consenting (65.5%) in an average of 1.2 days. Hybrid recruitment was used in 27.1% of profiles and resulted in a 44.5% consent rate in an average of 8.6 days, with electronic medical record messaging being the attempt type most likely to lead to a consent. Independent recruitment was used in less than 1% of profiles. In Active and Hybrid Recruitment, non-English speakers were more likely to consent compared with English speakers. Collectively, these findings emphasize that although optimal pilot NBS recruitment is multi-modal, direct communication between parents and study team has the highest consent yield.

## 1. Introduction

Newborn screening (NBS) is designed to detect serious, treatable conditions as early as possible, when therapy is likely to be most effective. The growth of biochemical, physiologic, and genetic screening technologies coupled with the expansion of novel genetic therapies has dramatically increased the urgency of assessing if and how to expand the scope of screenable disorders.^1^

Pilot NBS studies are an optimal way to collect objective information about the feasibility, net benefit, and acceptability of screening for specific conditions, which can inform decision-making about the appropriateness of inclusion of the condition(s) into public health-based NBS programs.^2^ Pilot studies typically involve screening a subset of newborns for a particular condition and are often used to evaluate the accuracy of screening tests, refine cutoffs, and to assess the impact of early detection on patient outcomes. Because of the rarity of most candidate NBS disorders, it is important that pilot programs be conducted in as large a population as feasible to increase the likelihood of detecting affected infants.

Some pilot NBS studies are de-identified and retrospective, using residual dried blood spots (DBS) to study assay accuracy, cutoff values, and disease incidence. This relatively inexpensive study design can provide fast, useful data about the feasibility of screening for a particular disorder in a public health lab.^3 4 5,6^However, with de-identified studies, the ability to compare screening results with clinical diagnosis is not possible, and abnormal results cannot be shared with families.

Identified studies, on the other hand, link the DBS sample with the infant. This provides critical feedback to the screening laboratory about confirmatory test results and offers the potential for a direct benefit by enabling clinical follow up of babies identified to be at risk. Some identified prospective pilot studies have used opt-out consent models, enrolling all infants unless their parents explicitly decline participation. ^2,7 8^Other pilot studies have been mandated at the state level, without requiring parental permission.^2 9^

In contrast, many identified prospective pilot studies require an informed consent process and only enroll infants whose parents opt in to participate. This approach may be taken based on the complexity of the screening panel, the inclusion of other research procedures like parent surveys, and the targeted patient population. While the benefits of this approach are vast, there are significant logistical, financial, and organizational challenges concerning the best methods to address large-scale parental recruitment and support informed decision-making. As a result, different studies have utilized recruitment processes that vary by timing, outreach, and communication modes. For example, the UK-based Generation Study starts recruitment in the prenatal period.^10^ Compared with recruiting during the postnatal period with its inherent demands on parental attention and time, prenatal recruitment models let parents learn about the study when they may have fewer demands on their attention. While this seems optimal and is supported by parental feedback, ^6^ large scale recruitment in the prenatal period is logistically challenging because of the wide scope and heterogeneous nature of obstetric settings. Thus, pilot study recruitment in the postnatal period is a more commonly used approach, as it enables outreach in a more uniform environment. Several published pilot studies have used primarily postnatal, in person recruitment where recruiters reach out directly to parents on the postpartum wards in the days immediately following the birth of their child. ^11 12,13^ While the Early Check study also recruited mainly in the postnatal period, it used a wide range of outreach modalities to target parents statewide, including mailed letters, patient portal invitations, social media advertisements, and community distribution of materials.^14,15 16^

These varying recruitment approaches have shown a similarly varied range of infant enrollment. The EarlyCheck study enrolled 12,065 infants between 2018–2020 reflecting 5% of state births.^16^ The first BabySeq study enrolled less than 10% of babies whose parents were approached to participate.^11^ Recruiters in a New York-based DMD pilot approached about half of all eligible families during its first year; of these, 85% enrolled their infants. ^12^ The ongoing GUARDIAN study also uses a predominantly postnatal, in person approach and has enrolled about 72% of approached families,^17^ similar to the 73% consent rate in our lysosomal disorder pilot screen that recruited in a similar fashion.^18^ In a prospective pilot screen for metachromatic leukodystrophy in Tuscany that required written consent, less than 0.2% of parents declined participation.^13^

These examples demonstrate the challenges of designing large scale consented newborn screening pilot studies in a way that adequately addresses parental education^19^ and engagement while working within infrastructural constraints, personnel limitations, and limited study budgets. Although there is likely no “one size fits all” approach, a comparison of the effectiveness of recruitment approaches may help inform future protocol development. While comparing recruitment efficacy across different studies is difficult given the inherent differences in piloted disorders, patient populations, and study design, doing so within the context of one large study may provide informative data.

ScreenPlus is a consented NBS pilot program that aims to enroll 100,000 babies from eight high birth rate pilot hospitals across New York City within a nine-year period. Using DBS already obtained for routine NBS, consented samples are screened at the New York State Newborn Screening Program using a multi-tiered approach for a panel of genetic disorders that have a pediatric phenotype, an approved therapy or open clinical trial, and a low cost, validated screening assay.^20^ Given the large-scale nature of this pilot program, ScreenPlus utilizes a variety of recruitment tactics to introduce parents to the study. In this paper, we take a deep analytical look at our recruitment strategies to determine the most effective and efficient modes that garner an informed decision from eligible families by evaluating the most common modes used at time of decision for parents/guardians, the number of attempts it takes a research coordinator before getting a final decision from families, and other factors that correlate with decision-making. We also explore the most common reasons parents decide to consent, as well as their feedback about the overall recruitment process.

## 2. Materials and Methods

### 2.1. Study Subjects

The inclusion criteria for participation include infant age between 0-34 days and being born at one of the ScreenPlus pilot hospitals. There are no exclusion criteria related to infant’s health, gestational age, or biological sex.

The study was approved by the single Institutional Review Board for the project, the Biomedical Research Alliance of New York (BRANY; Protocol #19-10-212). It is registered on clinicaltrials.gov(NCT05368038).

### 2.2. Pilot Hospitals

Recruitment for ScreenPlus is conducted at eight pilot hospitals in the New York City metropolitan area: Jack D. Weiler/Montefiore Einstein (Bronx), NYU Langone - Tisch Hospital (Manhattan), NYU Langone Brooklyn Hospital (Brooklyn), Maimonides Medical Center (Brooklyn), Long Island Jewish Medical Center, Northwell Health (Queens), North Shore University Hospital, Northwell Health (Long Island), Mount Sinai Medical Center (Manhattan) and Mount Sinai West (Manhattan). These sites were chosen because of high annual birth rates ranging from ∼3500 to 9000 per year, presence of expertise in neonatology and/or medical genetics, and their locations representing broad swaths of the NYC population. Each site has one full-time Research Coordinator (RC), several of whom are multilingual, to facilitate recruitment, site-specific regulatory processes, and data entry. Each site also has a Principal Investigator who is either a medical geneticist or neonatologist.

### 2.3. Recruitment Timeline

Recruitment began at the Jack D. Weiler Hospital in the Bronx in May 2021, despite ongoing waves of COVID-19 and resulting limitations on non-essential face-to-face interactions. The other pilot hospitals went live from August 2022 to November 2023, after sponsor contracting, site-specific sub-contracting, hiring, training, translations, and IRB approvals were completed. After recruitment began, most hospitals had one or more transient halts due to personnel turnover that lasted between three and nine months. The Independent (passive) recruitment database became available to parents through QR codes on ScreenPlus materials in October 2022. Recruitment is ongoing and is anticipated to continue through 2029.

### 2.4. Recruitment Materials

IRB-approved RC scripts (in-person discussions and phone calls), messages (email, text, electronic medical record (EMR)) and materials (brochures, welcome/discharge packet letters, posters) are used for recruitment. Materials are distributed and displayed in a variety of settings visible to parents, including maternity wards, labor & delivery floors, NICUs, and prenatal care provider offices. All materials, including the consent and HIPAA forms, are available in the ten most common languages spoken at the pilot hospitals, including English, Spanish, Simplified Chinese, Arabic, Hindi, Bengali, Urdu, Albanian, French and Russian.

### 2.5. Recruitment Modes

For **Active recruitment**, RCs initiate contact with the parents at the bedside in the postpartum unit. Active recruitment occurs before discharge, typically within 48 hours of the baby’s delivery. As each pilot site has only one full-time RC, this mode of recruitment is limited to the times when the RC is on site, generally weekdays during business hours. RCs use the EMR to obtain lists of daily inpatient postpartum mothers. They then approach each family to introduce them to ScreenPlus. As postpartum mothers may be unavailable for a variety of reasons (ex. resting, medical needs, receiving visitors), the RCs often make one or more attempts to meet with the family which may occur over one or more days. The ScreenPlus REDCap database captures the dates and outcomes of up to nine attempts to speak with the parents, preferred language based on the EMR, actual language(s) used for interaction, use of translators, and any additional notes that the RCs deem relevant. Attempt outcomes (consented, declined, no decision) are then documented.

**Hybrid recruitment** modalities are also RC initiated. They include a first attempt that is not in person (ex. initial contact made by phone call after discharge), or any combination of in-person, email, phone, text, and EMR messages; RC’s can choose which attempt type to use in any given situation. Hybrid recruitment often continues after discharge, and parents can ultimately decide to consent directly with the RC or do it on their own online using the eConsent. Similar to the data collected for Active recruitment, the RCs record all attempts, attempt type, and outcomes, as well as language.

For **Independent recruitment**, the parent initiates contact to participate in ScreenPlus. All the recruitment materials contain QR codes that bring interested parents to the REDCap-based multi-lingual passive recruitment forms. Initially, the parents fill out an eligibility form that inquires about the infant’s age and hospital of birth. If the infant is eligible, the parents are then directed to the e-consent that they can use for decision-making about study participation. Contact information for the RCs is available should the parents desire assistance or additional information.

### 2.6. Recruitment Outcomes

Successfully Approached refers to profiles with one or more documented attempts and a final outcome of **Consented, Declined, No Decision, or Ineligible**. Consented parents make the decision to participate via any of the recruitment modes noted above. Declined refers to situations where parents decide that they do not want to participate. No Decision refers to situations where contact is made between the RC and parents, but the parent did not provide a final consent or decline decision. These profiles are considered “decline by default” because the baby ages out of eligibility (post 34 days of age) without a documented decision. Ineligible refers to the rare situation where the infant was deemed ineligible after parental consent, typically due to prolonged recruitment time during which the infant aged out of the study.

Failed Approach refers to situations where there were attempts from the RC to engage with the parents, but no interactions took place. Common scenarios for Failed Approach include parents being unavailable/busy when the RC visits or calls, or not responding to an email, EMR message, or text message.

### 2.7. Surveys

At the end of recruitment encounters, parents are offered an opportunity to participate in anonymized feedback surveys about their reasons for consenting (“ELSI-C”) or declining (“ELSI-D”). ELSI-C aims to gain insights not only on why participants chose to consent for ScreenPlus, but also asks them about what study materials they saw and used, which were most useful, and how easy the electronic consent format was to use. ELSI-D focuses on parental concerns that might impact their decision to participate and is the subject of a future manuscript.

### 2.8. Data Capture and Statistical Analyses

A “profile” is defined as a single REDCap entry representing one mother/baby(ies) unit. Because recruitment decisions are made at a parent level, twins and triplets are counted as a single profile. As a result, the total number of profiles reflects the number of families rather than the total number of babies, thus underestimating the number of babies represented in the dataset. In cases where more than one REDCap profile existed for the families with multiple births, a single “master” profile was established for analysis based on which REDCap entry had the most consistently documented recruitment activity. The purpose of this process is to avoid double-counting and ensure complete capture of recruitment activity. Duplicate profiles were cross-checked to ensure all attempt-level data were consistent and accurately captured. Relevant recruitment information was integrated into the selected master profile used for analysis.

The analytic dataset included recruitment activity occurring between May 10, 2021, and April 15, 2025. Profiles were included only if all recruitment attempts and final outcomes occurred within this timeframe. Profiles were excluded if they contained any recruitment-related activity after April 15, 2025, where profiles had only recruitment related activity with an outcome of “Failed”, or if they had zero documented attempts and no accompanying contact notes or additional recruitment information. Any communications that were documented outside the timeframe that were unrelated to recruitment activities (e.g. relaying requested results or coordinating additional sample collection) were only excluded at the attempt level but did not result in exclusion of the overall profile, provided all recruitment-related activity occurred within the dataset time period. The exclusion process was applied consistently across all profiles before analysis.

Total estimated births during the recruitment period were derived from monthly counts of babies with at least one routine NBS specimen received by the New York State Newborn Screening Program as a proxy for total births at participating pilot hospitals. Counts were prorated for months with partial recruitment coverage, adjusted using a two-day lookback buffer at recruitment start dates, and were assigned a value of zero during periods without active RC coverage. Adjusted counts were summed to generate site-specific and overall birth estimates.

Recruitment time was calculated from the date of first contact to the date of final consent or decline, and for “No Decision” profiles, recruitment time was measured from the first attempted contact to the final documented attempt. Each in-person interaction, email, EMR message, phone call, and text message was counted as an individual attempt.

Data cleaning and finalization were conducted at the attempt level prior to profile-level analysis. Recruitment notes were systematically reviewed to identify any undocumented attempts or missing information regarding attempt mode and outcome. Missing information that was documented in open-text notes was incorporated into structured fields, when applicable (e.g. notes including text ‘left a voicemail’ or ‘EMR message sent’). The Independent database was exported and reviewed against the primary dataset to verify profile completeness and final outcomes. Duplicate passive entries were excluded to ensure each profile had one corresponding passive record.

Additional quality checks were applied to ensure consistency between recruitment attempts and recruitment outcomes. Following data cleaning, the attempts within each profile were organized chronologically to represent the order of attempts, duration of recruitment, and progression toward the recruitment outcome for this analysis.

## 3. Results

All data were analyzed within the period of recruitment from May 2021 through April 2025. An estimated 99,677 births occurred at participating pilot hospitals during the recruitment window and were used as a proxy for total eligible births.

The recruitment process was initiated with 58,545 families, representing 58.7% of eligible births. Approximately 41,132 babies born at the pilot hospitals during the recruitment period were not approached by RCs, reflecting the “weekday only” limitations of our protocol, which excluded RC-initiated recruitment on weekends, sick days, vacation days, and other periods when RCs were unavailable.

Figure 1 summarizes recruitment modes used for 58,545 approached families. Of these, 10,903 resulted in failed approaches, where RCs attempted to approach families but were unable to make contact. The remaining 47,642 families were successfully approached and comprise the study population included in downstream analyses. Among these families, Active recruitment was the most used mode (72.2%, 34,411), followed by Hybrid recruitment (27.1%, 12,918). Parent-initiated Independent recruitment accounted for less than 1% of completed profiles (313).

**Figure 1.**
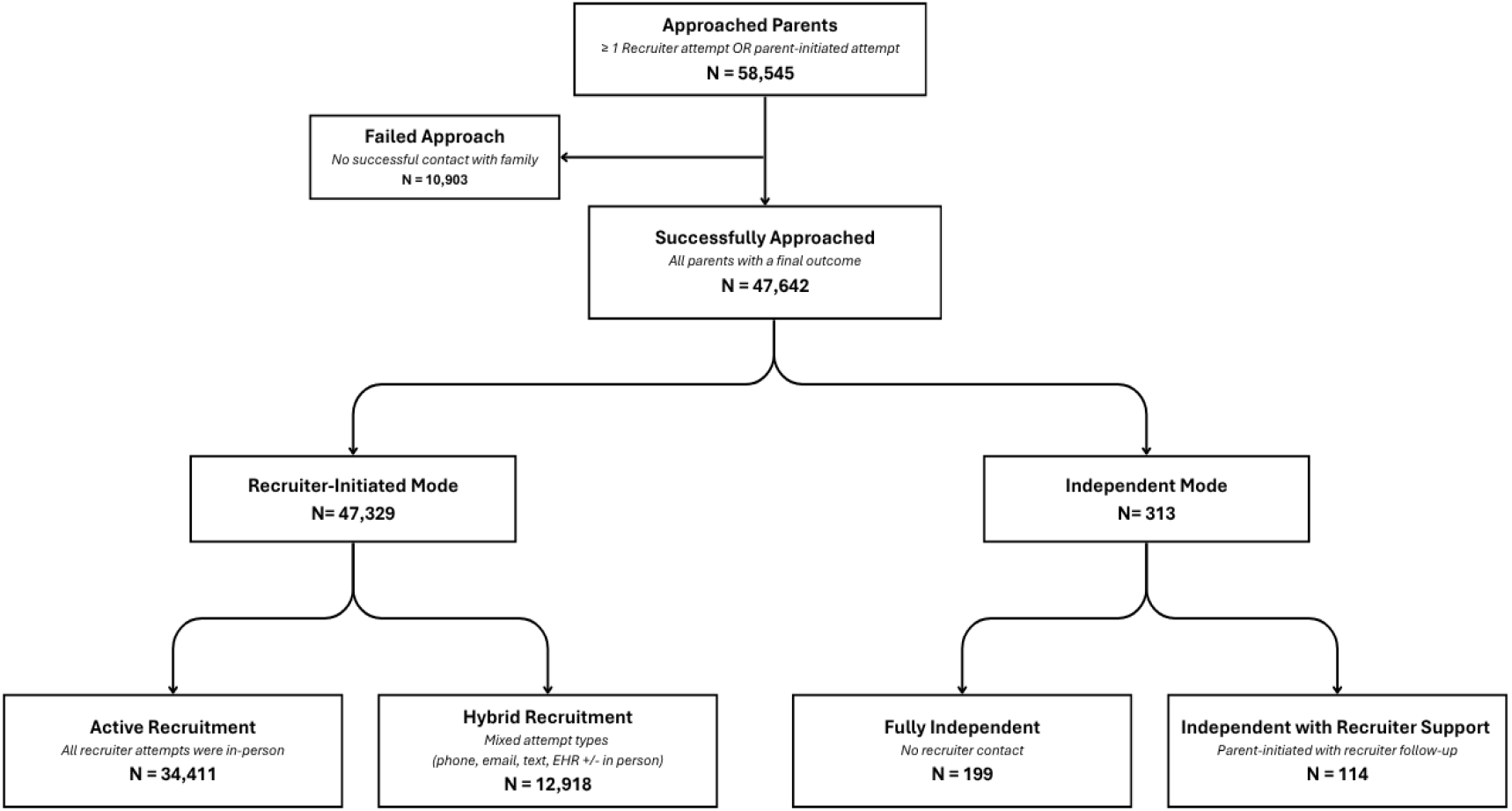
Overview of Recruitment Modes. A summary of the downstream of “successfully approached” parents into the various recruitment modes.

When considering overall outcomes among successfully approached parents regardless of recruitment mode, 59.9% (28,558/47,642) consented to participate, 18.9% (9,001/47,642) declined, 21.1% (10,057/47,642) made no decision, and 0.05% (26/47,642) were ineligible. Relative to the estimated 99,677 births occurring at participating hospitals during the study period, 28.7% (28,558/99,677) were ultimately enrolled in ScreenPlus.

Figure 2 shows final outcomes among the 47,642 parents who were successfully approached. The distribution of recruitment outcomes varied by recruitment modes, as detailed below in Figure 2.

**Figure 2.**
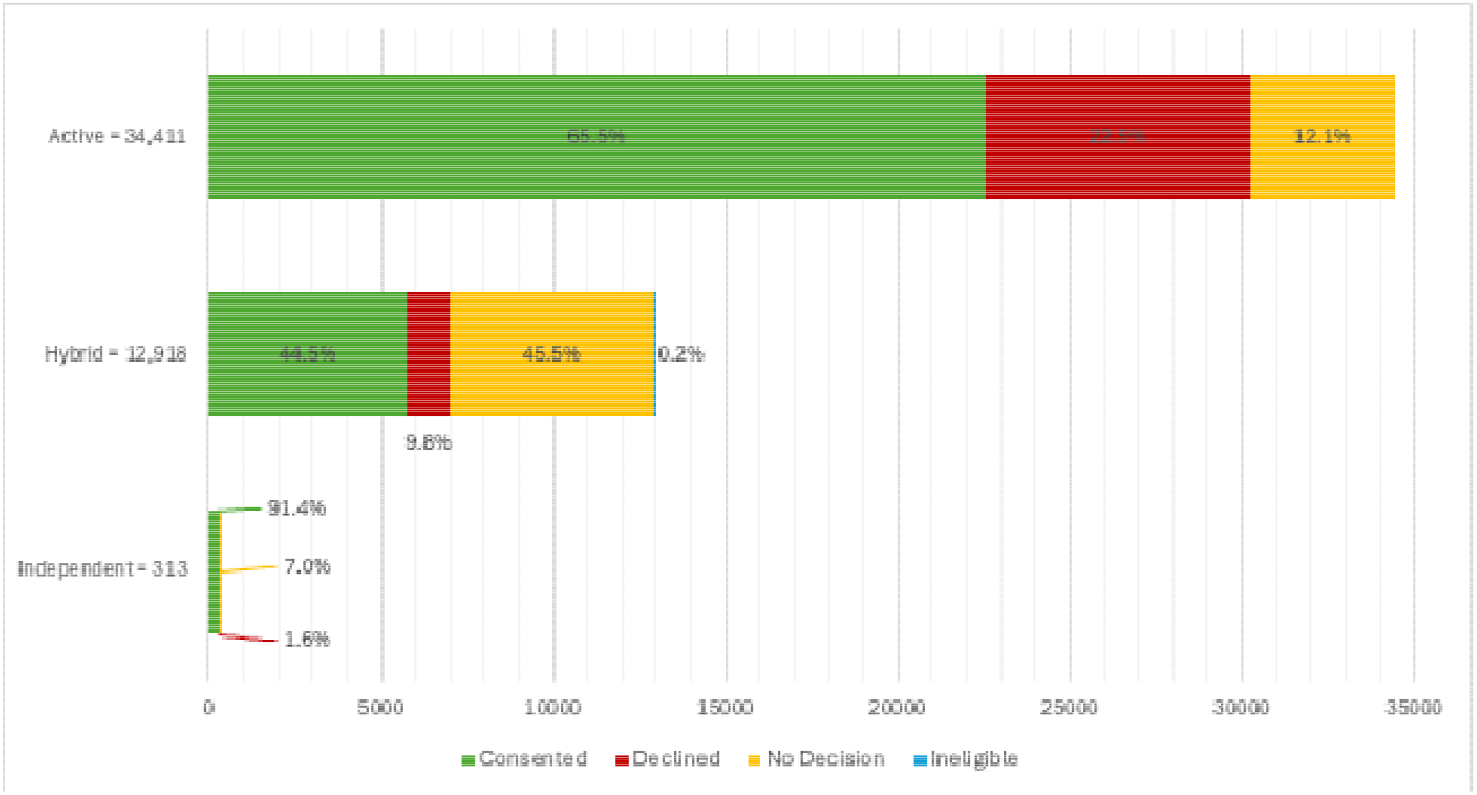
Distribution of final outcomes among successfully approached parents by recruitment mode (N=47,642). Active recruitment reflects in-person recruiter-initiated contact only; Hybrid recruitment includes mixed contact types; and Independent recruitment reflects parent-initiated. Sample sizes (34,411, 12,918, 313) are shown for each recruitment mode.

### 3.1. Active Recruitment

Of the 34,411 parents recruited through Active recruitment mode, which consisted only of in-person interactions with RCs, 22,526 (65.5%) consented, 7,733 (22.5%) declined, and 4,152 (12.1%) made no decision (Fig. 2). Among parents who made a consent or decline decision, 95.4% (28,865/30,259) did so within the first two attempts, with relatively few decisions made after the third attempt (Fig. 3). Overall, the average recruitment time was 1.2 days (range 1-35) from the date of initial attempt to the date of final outcome or final attempt, and the average number of attempts was 1.3 (range 1-6).

**Figure 3.**
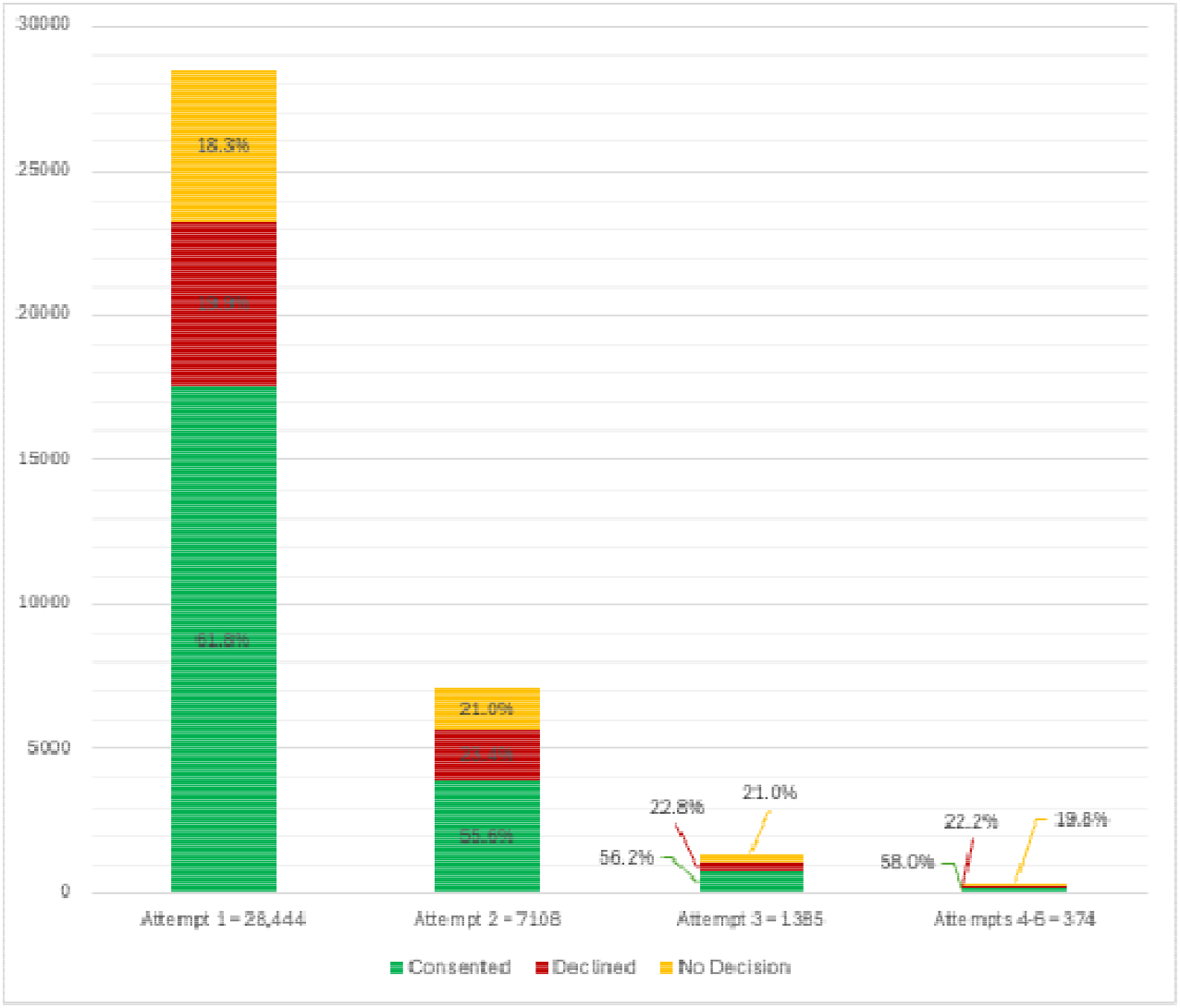
Active recruitment final outcomes (consent, decline, no decision) by attempt number (N=37,311). The N for this graph is larger than the overall N of the group (34,411) as those who did not consent/decline in the first attempt, had a first attempt outcome of no decision, thus had multiple attempts.

### 3.2. Hybrid Recruitment

Of the 12,918 parents in Hybrid recruitment, 5,746 (44.5%) consented, 1,263 (9.8%) declined, 5,883 (45.5%) made no decision, and 26 (0.2%) were ineligible (Fig. 2). As Hybrid recruitment often continued after hospital discharge, the average recruitment time was 8.6 days (range 1-233), with an average of 2.8 attempts (range 1-9). For parents who consented, the average recruitment time was 5.0 days and 2.6 attempts, while parents who declined required on average 9.3 days, and 2.6 attempts.

When evaluating the attempt number at which a decision was made, 11.1% (636/5,746) of consenting parents decided on the first attempt, 47.4% (2,723/5,746) on the second attempt, and 25.1% (1,441/5,746) on the third attempt (Fig. 4a). Consent frequency decreased with subsequent attempts. Among parents who declined, 14.7% (186 /1,263) did so on the first attempt, 39.8% (503/1,263) on the second attempt, 27.0% (341/1,263) on the third attempt, and 18.4% (233/1,263) on subsequent attempts (Fig. 4b).

**Fig. 4.**
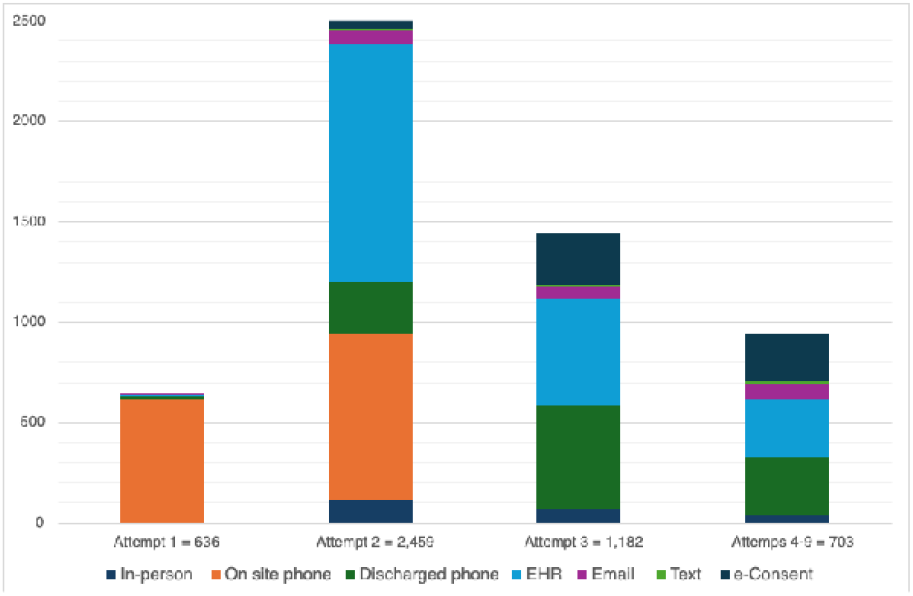
Hybrid recruitment summary. Within each figure, the bars represent the attempt number at which the final outcome was made and are further categorized by the attempt type used.

**Figure 4a.**
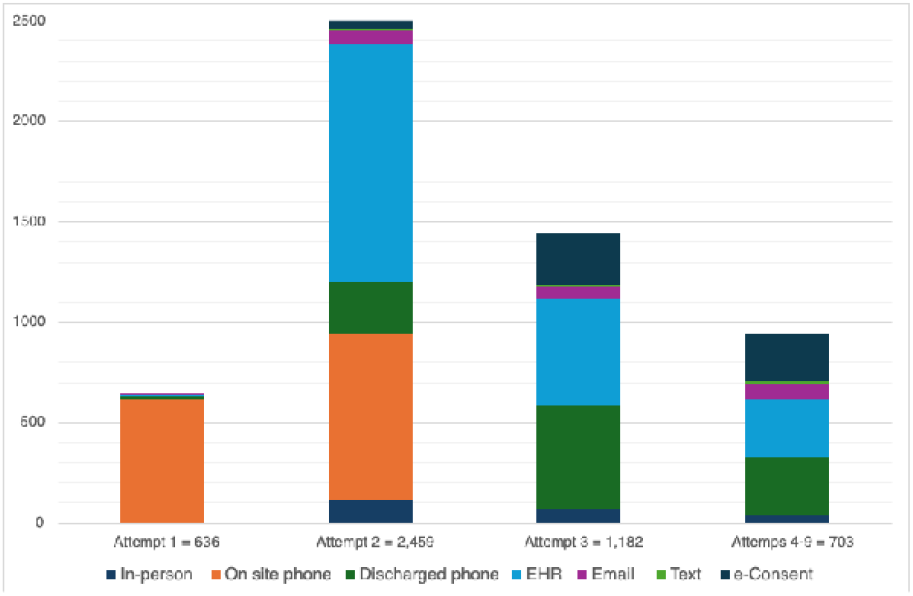
Attempt types that resulted in parental consent.

**Figure 4b.**
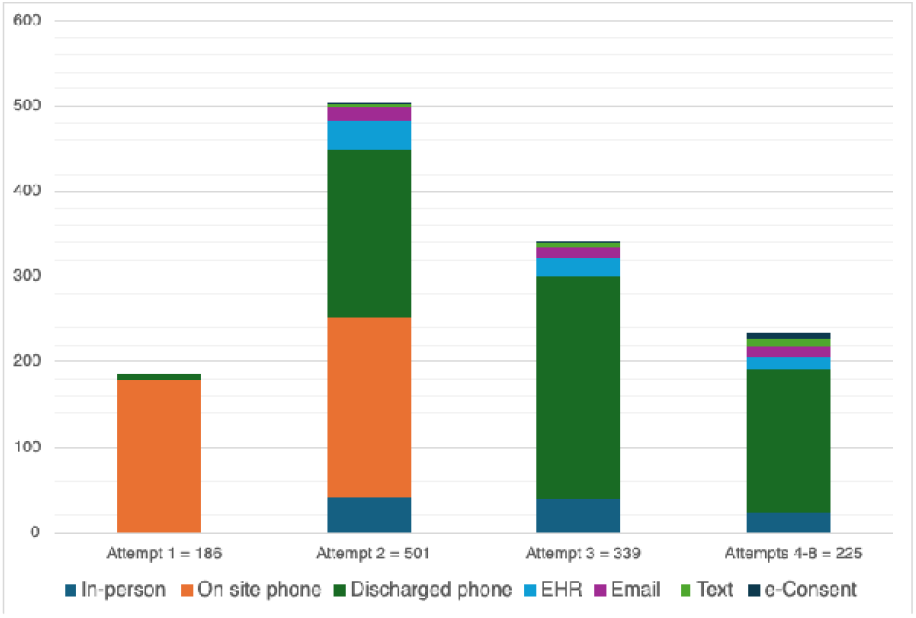
Attempt types that resulted in parental decline.

Examining the attempt types used when a consent decision was made, EMR messaging was the most frequent type (34.9%; 2,007/5,746), followed by onsite phone calls (25.0%; 1,437/5,746), post-discharge phone calls (18.9%; 1,088/5,746), and e-consent (13.3%; 766/5,746) (Fig. 4a). Email and text messages were infrequently used. Parents who declined did so mainly by phone, post-discharge (50.4%, 636/1,263) or onsite (30.6%, 386/1,263) (Fig. 4b).

### 3.3. Independent Recruitment

A total of 313 parents entered the Independent recruitment mode through parent-initiated contact. Of these, 286 (91.4%) consented, 5 (1.6%) declined, and 22 (7.0%) did not complete the consent process.

Among the 286 parents that consented, 87 (30.4%) required RC follow-up support, while 199 (69.6%) completed the consent process independently. For those needing RC support, the average time to consent was 12.5 days (1-31) and required an average of 2.7 attempts (1-6). Email was the most used follow-up type, followed by EMR messaging.

### 3.4. Site differences

Despite the use of a uniform protocol and recruitment materials, variation in recruitment outcomes was seen across pilot sites (Table A2). The proportion of estimated births that were successfully approached ranged from 24.2% to 73.6% across sites. Differences were also seen in the use of Hybrid recruitment attempt types across sites (Figure A1), including variation in the relative use of in-person, phone calls, EMR, email, and text messaging.

### 3.5 Language and demographics

Languages used during recruitment are shown in Table 1. Among successfully approached families, 86.2% preferred English, 7.4% Spanish, and 6.4% another language. The language distribution was similar across Active and Hybrid recruitment modes.

**Table 1.**
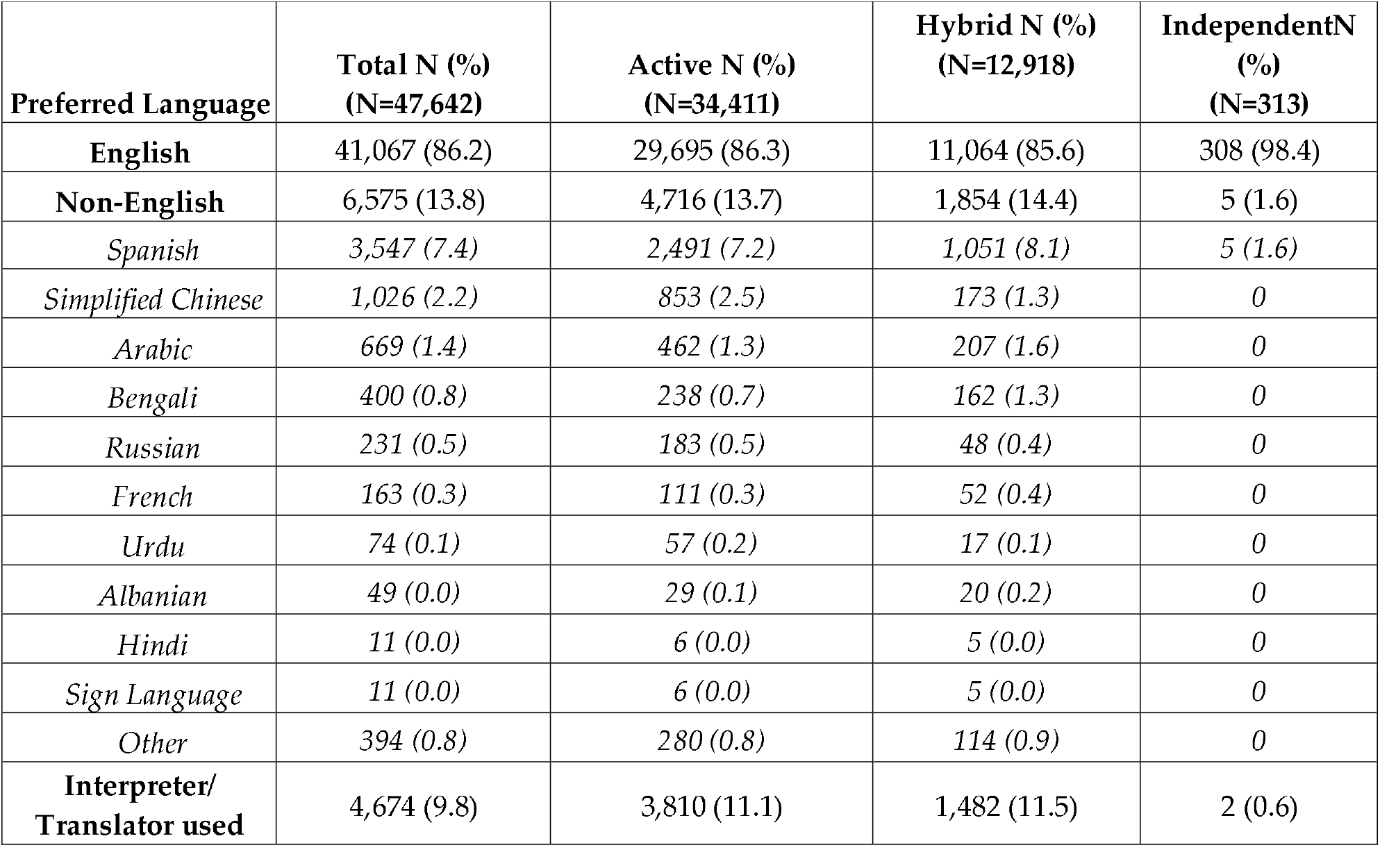
Preferred language and translator use among successfully approached ScreenPlus families by recruitment mode.

Consent rates differed by preferred language and recruitment mode. In Active recruitment, 63.8% (18,933/29,695) of English-speaking parents consented compared with 76.2% (3,593/4,716) of parents with a preferred non-English language. In Hybrid recruitment, 43.2% (4,779/11,064) of English-speaking parents and 52.2% (967/1,854) of non-English parents consented. Consent rates varied across specific languages, ranging from 16.7% in American Sign language to 86.5% for Simplified Chinese (data not shown). This will be further elaborated in a future manuscript.

Race and ethnicity data are not routinely collected as part of routine NBS in New York and were therefore not collected during recruitment for ScreenPlus. Self-reported demographic data is collected through ScreenPlus surveys, including those taken at the time of consent (ELSI-C) and serves as a proxy for overall ScreenPlus recruitment demographics (Table A1).

### 3.6. Parental feedback

Of the 28,558 consented parents, 2,486 (8.7%) completed the consenter feedback survey (“ELSI-C”). For the survey item assessing which materials were used to learn about ScreenPlus (n=2,460), 1,604 (65.2%) parents reported learning about ScreenPlus through “Discussions with the Research Coordinator.” When asked which resource was the most helpful for making the decision to participate (n=2,466), 1,585 (64.3%) found those discussions to be the most useful educational resource. The e-consent and ScreenPlus website were less frequently selected as the most useful educational resource, with 317 (12.9%) and 150 (6.1%) parents selecting these options, respectively.

Reasons for consenting are shown in Figure 5. Parents most commonly cited obtaining information about their baby’s health (46.0%) and participating in research that might help other babies (41.3%). When asked about the ease of the study’s e-consent process, 764 parents responded, with 753 (98.6%) reporting that the format was “very easy” or “easy” to use.

**Figure 5.**
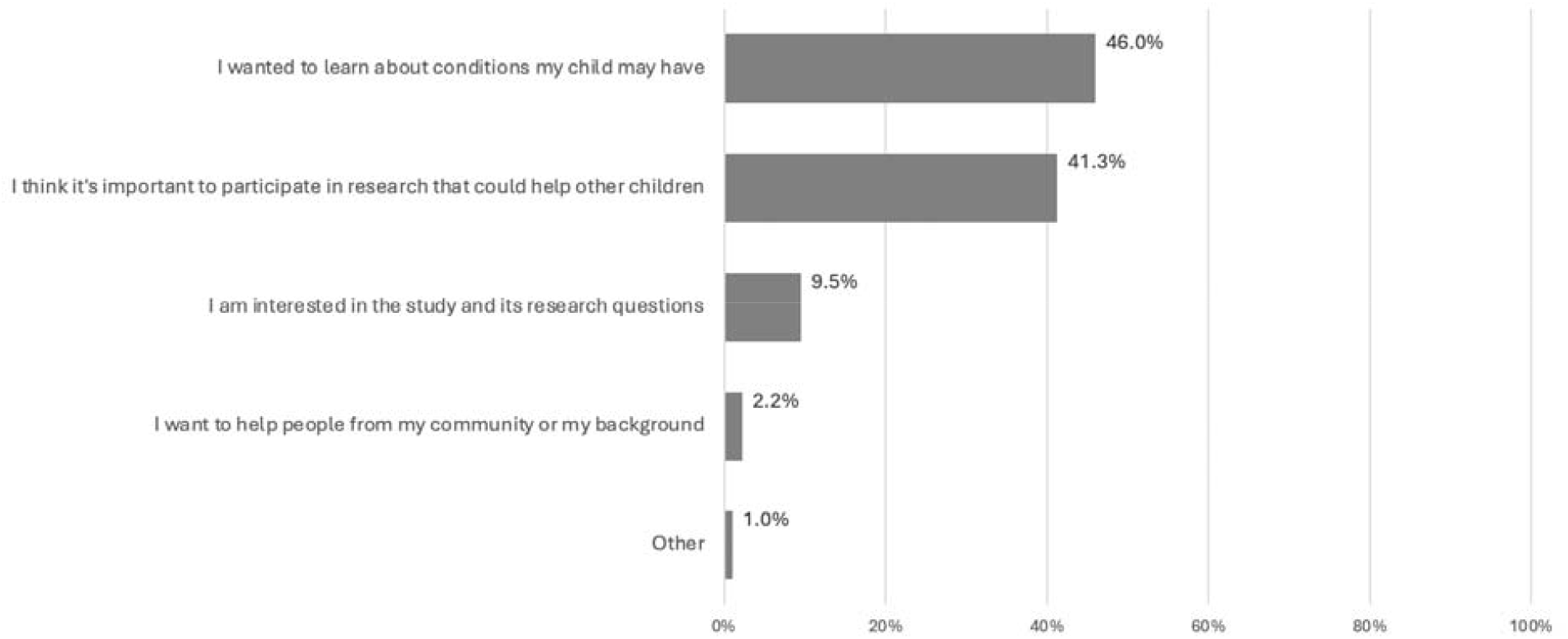
Distribution of parent responses to the survey question assessing the main reason for participating in ScreenPlus (n= 2,476). Missing responses were excluded from the analysis (n= 10); percentages are based on the respondents who answered this survey item.

## 4. Discussion

Pilot NBS studies play a critical role in the expansion of NBS by supplying objective data about the feasibility, accuracy, and outcome of additional screening. Pilot studies are now even more crucial as NBS is evolving to consider the use of genome sequencing as a screening tool. This transformation requires the ability to design trials that collect objective feasibility and outcome data. Equally important, however, is the ability to design trials that emphasize parental education and informed decision-making.

One of the major challenges in designing pilot NBS studies is striking the balance between enrolling the large numbers of infants needed for scientific validity and optimizing recruitment/informed consent processes to ensure that parents have enough time and information to make a meaningful decision about participation. On one hand, focusing primarily on enrollment numbers may yield important screening data but can come at the expense of parental engagement and education, leading to consent that is not truly “informed.” On the other hand, recruitment/informed consent models that are overly complicated and lengthy may be difficult to implement in a busy hospital setting. They may also cause parents to shy away from participation because of the perceived complexity and risks, leading to low enrollment numbers that may not provide enough data about screening outcomes.

Many pilot NBS programs have been thoughtfully developed with these issues in mind. However, additional factors must also be taken into account, such as time limits of the program, limited research budget, target population, and systemic infrastructure. For these reasons, pilot programs frequently differ in design. The EarlyCheck program, for example, targeted all babies born in North Carolina so developed a statewide extensive multimodal outreach program to introduce parents to the study and enable them to self-enroll.^15^ Other studies, such as BabySeq and GUARDIAN,^11 17^have focused on recruiting infants within specific hospital systems, making in-person recruitment feasible. Given these differences in study design, it is difficult to compare the effectiveness of recruitment strategies across different pilot programs.

One of the strengths of this ScreenPlus recruitment data analysis is the underlying uniformity of the disorder panel, processes, materials, and timeframe, which enables a cleaner comparison of the efficacy of different approaches. The slight differences in approaches and enrollment numbers between sites were expected and may reflect the unique environment of each hospital system. It also demonstrates the importance of conducting pilot NBS programs in several sites to reduce the risk of artificially skewed data.

Overall, the data clearly shows the superiority of Active, in-person recruitment at eliciting a parental decision. Most parents who consented to participate did so after discussions with a RC. When a decision was not made within the first or second attempt, the likelihood that a parent agreed to participate fell with each subsequent attempt. This data suggests the optimal number of attempts is two, with diminishing return after three attempts. Therefore, a study design that limits the number of in-person recruitment attempts may optimize RC time and effort.

While effective, Active recruitment at the bedside in the postnatal period carries with it the inherent challenges of engaging with new parents in the days after the arrival of their baby. Medical concerns, fatigue, medications, visitors, and other competing interests can make this a less-than-ideal situation for study discussions. Because of this, ScreenPlus RCs can also use a more flexible Hybrid recruitment mode that enables them to interact with parents over a longer period using mixed outreach types, including calls, text messages, emails, and EMR messages. Interestingly, EMR messages from RC to parents resulted in a relatively higher consent rate compared with other attempt types, suggesting that this may be an efficient way to engage parents. Hybrid recruitment often continues after patient discharge. As a result, parents recruited through Hybrid recruitment take longer on average to make a decision than parents recruited through Active in-person recruitment. In addition, the consent rate is relatively lower, and the “no decision” rate is higher. However, the breadth of Attempt types that were effectively used suggests that flexibility remains an important Recruitment tool.

Parent–initiated Independent recruitment seems conceptually the easiest and least complicated way to engage parents. Our Independent scheme relied on the display of study posters and brochures in high-visibility clinical areas as well as inclusion of study materials in hospital welcome and/or discharge packets. Interested parents could initiate recruitment by accessing eligibility and e-consent forms using QR codes. The number of parents who consented via the Independent mode was surprisingly low, with only 286 parents (1.0% of total consented) enrolling this way. Furthermore, 77 additional parents (not included in the analytic population) were noted to have started the passive eligibility and/or consent forms but were not enrolled. Of these, 58 (75.3%) were missing critical information needed to determine eligibility (ex. names, date of birth), 18 (23.4%) were eligible but did not complete the e-consent form, and one was ineligible due to age out. This may reflect a suboptimal study material dissemination plan that did not include targeted social media outreach as a conduit to enrollment, an approach that was used in the EarlyCheck study.^15^

Collectively, these findings emphasize that RC-initiated face-to-face contact significantly drives parent decisions and informed discussions, a concept that was reinforced by parental surveys showing that in-person discussions were the most useful source of information. This likely reflects the importance of human interaction at enhancing trust, clarity, and perceived legitimacy of the study. However, pilot studies should be designed to allow for flexible paths to enrollment. Flexible approaches, such as our Hybrid model, may be less efficient in terms of time and effort to elicit a parental decision, but are still important to optimize equitability and outreach. Patient-initiated Independent recruitment yielded the highest percentage of consent but was used by a small fraction of participants. This may reflect inadequate dissemination of study information and highlights the need to study if expansive targeted outreach efforts can enhance the use of passive recruitment in large pilot programs.

Of note, ScreenPlus focuses on recruitment during the postnatal period. This decision was made because the large numbers of prenatal clinical programs associated with our pilot hospitals made in-person recruitment during this period logistically and financially unfeasible. Experts have recommended study introduction in the prenatal period, when parents presumably have more time to learn and decide.^2,6^ Designing pilot programs that can effectively engage parents during prenatal visits is therefore desirable, although the challenge persists on how to best incorporate high visibility study recruitment materials and personnel across the large number and wide range of obstetric clinical programs. Thus, this is an area requiring further exploration.

Important concerns about disparities in pilot study participation remain. Some studies have shown that minority and lower-income families are less likely to be recruited, often due to mistrust, lack of access, or confusion about how pilot studies differ from routine NBS.^21^ This may, in part, reflect language barriers. Interestingly, we found that during both Active and Hybrid recruitment, relatively higher percentages of non-English speakers chose to consent compared to English speakers. Although this may be specific to our multi-cultural New York City population where the use of translators is routine and common, it does need further investigation, particularly in different regions and populations.

It is interesting to consider parental feedback about recruitment and engagement preferences in pilot NBS programs. In one study, parents expressed preference for a mixed approach that includes receiving information from trusted healthcare providers but enrolling independently online.^14^ In contrast, ScreenPlus parents reported that the most useful materials to guide their recruitment decision was direct interaction with our RCs, with relatively few finding the online material helpful. Although this may be specific to our study, there is always concern that online or remote Independent recruitment may result in unintended inequities in enrollment, particularly among parents with less familiarity or access to technology. Thus, large pilot programs like the Generation Study and BEACONS-NBS that use multi-modal recruitment methods in populations with wide ranges of socio-economic and geographic diversity may help inform about how to use recruitment materials and modes to enhance equitable access to participation.

In summary, these data reinforce that large scale recruitment in pilot NBS is most effective when utilizing direct interactions between RC and parents, and that most parents who consent do so within the first two attempts. When more flexible pathways are used, EMR messaging is more effective compared with other attempt types, followed by phone calls. Parent-initiated Independent recruitment is the least effective recruitment mode, but its efficacy requires further study using more extensive and creative outreach. Overall, these findings provide guidance to help design future pilot NBS studies in a manner that optimizes parental engagement and study efficiency.

## Data Availability

The datasets presented in this article are not readily available because the data are part of an ongoing study. Requests to access the datasets should be directed to the corresponding author.

## Embedded Tables and Figures

Fig 1: Overview of Recruitment

Fig 2: Decision Outcomes

Fig 3: Active Recruitment: Decision Outcome by Attempt Number

Fig 4a-b: Hybrid Recruitment: Attempt Type by Attempt Number

Fig 5: ELSI-C Results

Table 1: ScreenPlus Languages

## Appendix A

Table A1: ELSI-C Demographics

Table A2: Pilot Site Characteristics

Figure A1: Pilot Site Differences in Hybrid Recruitment Attempt Type

## Author Contributions

Conceptualization: MC, KP, NK, AG, and MW; Methodology: MC, KP, NK, AG, and MW, Software: MC, KP, and NK; Validation: MC, KP, and NK; Formal Analysis: MC, KP, NK, AG, and MW; Investigation: MC, KP, NK, SB, MJ, GK, RL, JG, AS, JO, AG, and MW; Data Curation: MC, KP, NK, AG, and MW; Writing-Original Draft Preparation: MC, KP, NK, and MW; Writing-Review and Editing: MC, KP, NK, SB, MJ, GK, RL, JG, AS, JO, AG, and MW; Visualization: MC, KP, NK, and MW, Supervision: MC, KP, NK, and MW, Project Administration: NK, KP, MC; Funding Acquisition: MW, NK

## Institutional Review Board Statement

The study was approved by the single Institutional Review Board for the project, the Biomedical Research Alliance of New York (BRANY; Protocol #19-10-212).

## Informed Consent Statement

Informed Consent was obtained from all subjects who participated in ScreenPlus

## Acknowledgements

Special thanks to our Research Coordinators: Kaylyn Abdul Jabar, Isabella Buitron, Rewaa Elgazzar, Anneke Fleming, Sabine Jean-Guillaume, Katherine Lambert, Mariam Mahgoub, Nahid Manzur, Aliza Quinones, Sylvia Scheiner, Benjamin Tamarin, Mian Hua Zheng

## Appendix A

**Table A1.** Demographic characteristics of ELSI-C sub-population (N = 2,486)

|  | N | % |
| --- | --- | --- |
| **Relation** |  |  |
| Mother | 2,276 | 91.6 |
| Father | 200 | 8.0 |
| Missing | 10 | 0.4 |
| **Age (in years)** |  |  |
| Under 18 | 19 | 0.8 |
| 18-24 | 155 | 6.2 |
| 25-34 | 1,244 | 50.0 |
| 35-44 | 1,014 | 40.8 |
| 45 and above | 35 | 1.4 |
| Prefer not to answer | 10 | 0.4 |
| Missing | 9 | 0.4 |
| **Education level** |  |  |
| High school degree or less | 357 | 14.4 |
| Some college | 184 | 7.4 |
| Trade/technical/vocational training or Associates degree | 129 | 5.2 |
| Bachelor's degree (e.g. BA, BS) | 730 | 29.4 |
| Master's degree (e.g. MA, MS) | 695 | 28.0 |
| Doctorate (e.g. PhD, EdD) | 311 | 12.5 |
| Other | 15 | 0.6 |
| Prefer not to answer | 60 | 2.4 |
| Missing | 5 | 0.2 |
| **Race/Ethnicity** |  |  |
| Non-Hispanic/Latino(a) Subgroups |  |  |
| American Indian, Native American, or Alaska Native, non-Hispanic/Latino(a) | 10 | 0.4 |
| Asian, non-Hispanic/Latino(a) | 369 | 14.8 |
| Black or African American, non-Hispanic/Latino(a) | 267 | 10.7 |
| Native Hawaiian/Pacific Islander, non-Hispanic/Latino(a) | 1 | 0.0 |
| White or European American, non-Hispanic/Latino(a) | 929 | 37.4 |
| Middle Eastern or North African/Mediterranean, non-Hispanic/Latino(a) | 42 | 1.7 |
| Multiracial, non-Hispanic/Latino(a) | 56 | 2.3 |
| Hispanic/Latino(a) Subgroups |  |  |
| Self-Selected Only (No other race), Hispanic/Latino(a) | 572 | 23.0 |
| American Indian, Native American, or Alaska Native, Hispanic/Latino(a) | 2 | 0.1 |
| Asian, Hispanic/Latino(a) | 4 | 0.2 |
| Black or African American, Hispanic/Latino(a) | 43 | 1.7 |
| Native Hawaiian/Pacific Islander, Hispanic/Latino(a) | 0 | 0.0 |
| White or European American, Hispanic/Latino(a) | 37 | 1.5 |
| Middle Eastern or North African/Mediterranean, Hispanic/Latino(a) | 1 | 0.0 |
| Multiracial, Hispanic/Latino(a) | 14 | 0.6 |
| Unknown | 26 | 1.0 |
| Prefer not to answer | 96 | 3.9 |
| Missing | 17 | 0.7 |
| **Total Household Income** |  |  |
| Less than \$49,999 | 465 | 18.7 |
| \$50,000 - \$99,000 | 299 | 12.0 |
| \$100,000-149,999 | 277 | 11.1 |
| More than \$150,000 | 954 | 38.4 |
| Don't know | 111 | 4.5 |
| Prefer not to answer | 367 | 14.8 |
| Missing | 13 | 0.5 |

**Table A2.** ScreenPlus pilot sites characteristics.

| Pilot sites | Recruitment start date | EPIC Platform Usage | Total Estimated Births | Successfully Approached (% of Estimated Births) | Total Consents | Total Declined | Total No Decision |
| --- | --- | --- | --- | --- | --- | --- | --- |
| Jack D. Weiler Hospital/ Montefiore | May 2021 | Yes | 11,294 | 8,314 (73.6%) | 4,119 | 802 | 3,391 |
| NYU Langone – Tisch Hospital | August 2022 | Yes | 12,497 | 6,351 (50.8%) | 4,624 | 1,113 | 607 |
| NYU Langone Hospital - Brooklyn | September 2022 | Yes | 11,012 | 6,409 (58.2%) | 4,852 | 1,044 | 512 |
| Mount Sinai Hospital | October 2022 | Yes | 16,053 | 8,890 (55.4%) | 4,585 | 2,501 | 1,802 |
| Mount Sinai West | October 2022 | Yes | 13,226 | 7,074 (53.5%) | 4,378 | 1,163 | 1,519 |
| Long Island Jewish Medical Center | January 2023 | No | 16,085 | 3,900 (24.2%) | 2,472 | 772 | 656 |
| Northshore University Hospital | February 2023 | No | 13,243 | 3,215 (24.3%) | 1,788 | 554 | 873 |
| Maimonides Medical Center | June 2023 | No | 6,267 | 3,489 (55.7%) | 1,740 | 1,052 | 697 |

**Figure A1.**
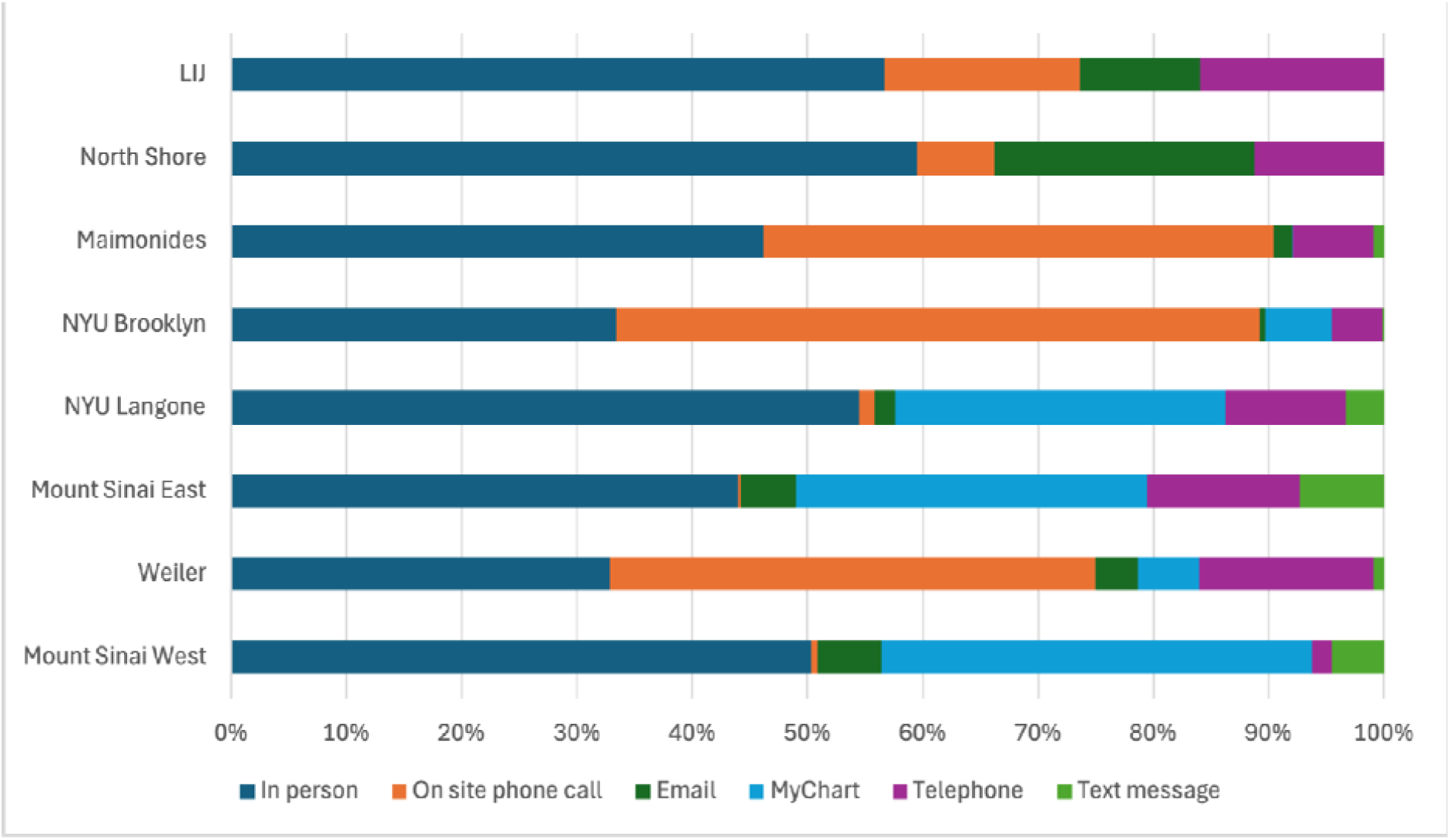
Attempt types utilized for Hybrid Recruitment by ScreenPlus pilot sites

